# A Bibliometric and Visualized Analysis of Transient Receptor Potential Vanilloid 1 in Allergic Diseases: Mapping the Knowledge Landscape

**DOI:** 10.1101/2025.09.21.25336288

**Authors:** Zhang Haoxiang, Wang Yanjie, Zhao Changqing, Cheng Fengli, Qi Xueping, Yang Junyue, Zhu XiaoJia, Wang Luyao, Shi Xiaoxi, Xu Danni, Fu Sirui, Duan Hongying

**Author notes:** Corresponding author: Zhao Changqing. Fund program: the National Natural Science Foundation of China (82171119, 82201263, 82301287); National Key Research and Development Program of the Ministry of Science and Technology of China(2023YFC2507905).

## Abstract

**Background:** Among the members of the Transient Receptor Potential (TRP) family, TRPV1 is distinguished as the primary receptor involved in thermal pain perception, closely associated with temperature changes. Acting as a critical nociceptor and thermosensitive receptor, TRPV1 is pivotal in the domain of allergic diseases. Despite its significance, there is a discernible lack of comprehensive reviews exploring the interactions between TRPV1 and allergic conditions in the academic literature. This study aims to thoroughly examine the relationship between TRPV1 and allergic diseases using bibliometric and visual analysis techniques. Our goal is to uncover the mechanisms by which TRPV1 influences allergic diseases, providing a robust scientific basis for future research directions and potential therapeutic strategies.

**Methods:** To investigate the relationship between TRPV1 and allergic diseases, we conducted a comprehensive search in the Web of Science (WoS) database. We employed a suite of analytical tools, including Excel, the WoS online analysis platform, the bibliometrix package in R, and CiteSpace software. These tools facilitated the systematic organization, detailed description, and thorough analysis of the retrieved literature, using co-citation, co-authorship, and co-occurrence visualizations to identify significant patterns and trends.

**Results:** This bibliometric analysis encompassed 1,045 articles on TRPV1 and allergic diseases, published by researchers from 57 countries and 1,179 institutions across 369 journals. Publication output remained low until 2008, after which it grew steadily, peaking at 70 articles in 2020. The United States contributed the most publications, followed by China and the United Kingdom. Johns Hopkins University emerged as a central collaborative hub, with Bradley Joel Undem as the most prolific author (30 publications). The most cited study, by Cevikbas et al., explored TRPV1/TRPA1 involvement in T-cell dependent itch. Recent research focuses on mechanisms such as neurogenic pain, itch, sensitization, oxidative stress, and atopic dermatitis, frequently employing mouse models.

**Conclusion:** This study provides a detailed analysis of research characteristics, emerging trends, and future directions in the study of TRPV1 and allergic diseases, offering an objective overview of global contributions in this field. It delivers critical insights to inform future investigations on TRP channels and allergic conditions. As an essential thermosensitive receptor, TRPV1 plays a central regulatory role in allergic diseases, with recent research increasingly directed towards developing animal models and clarifying activation mechanisms. Future research is expected to explore the complexities of sensitization mechanisms and receptor expression more deeply.Given its critical role, TRPV1 stands out as a promising pharmacological target for allergic rhinitis, meriting further study and potential therapeutic development. This focus on TRPV1 could lead to novel interventions that improve management and treatment outcomes for allergic conditions.

## Introduction

Recent research has highlighted the role of Transient Receptor Potential (TRP) channels, a class of transmembrane channel proteins found on the cell membrane. These channels are crucial for an organism’s ability to sense and respond to external environmental changes and stimuli. They are involved in numerous vital physiological processes, helping the body adapt and react to a wide range of physical and chemical stimuli. This fundamental role underscores the importance of TRP channels in maintaining homeostasis and contributing to the body’s overall functionality^[1]^.In 2021, scientists David Julius and Ardem Patapoutian were honored with the Nobel Prize in Physiology or Medicine for their pioneering work on the mechanisms of temperature sensation and touch perception. A significant focus of their research was on the TRP ion channel family, which acts as crucial sensors for detecting cold and heat stimuli. Currently, there are 28 TRP proteins identified in mammals, and these are organized into six subfamilies: TRPC (Canonical), TRPV (Vanilloid), TRPM (Melastatin), TRPA (Ankyrin), TRPML (Mucolipin), and TRPP (Polycystin)^[2]^. This classification underscores the diverse roles these channels play in sensory perception and various physiological processes.Among the ion channels most commonly implicated in thermosensation are TRPM8, TRPA1, and TRPV1. These channels function as principal detectors of environmental temperature extremes, sensitively responding to both cold and heat stimuli^[3]^.These three subtypes represent some of the most extensively researched and highly regarded ion channels within the TRP family. They are capable of detecting a variety of stimuli, including temperature, mechanical, and chemical signals, and play pivotal roles in processes such as analgesia, anti-pruritus, anti-inflammatory, and anti-edema^[4]^.

TRPV1, functioning as a pivotal thermosensitive and nociceptive receptor, plays an essential role in the modulation of pain and inflammation. Recently, it has garnered increasing attention in the research of allergic diseases^[5]^.AAs its name implies, TRPV1 is highly sensitive to capsaicin, which activates the channel through interaction with its binding site^[6-7]^.Caterina MJ and colleagues were the first to isolate a functional cDNA encoding the capsaicin receptor from sensory neurons, which they designated as VR1 (now known as TRPV1).This receptor is a polypeptide with six transmembrane domains, featuring a distinctive short hydrophobic region between the fifth and sixth transmembrane domains. The study identifies TRPV1 as an ion channel located at nerve terminals, predominantly expressed in nociceptive neurons. It functions as a critical molecular entity for sensing heat and pain, becoming activated at temperatures exceeding 43°C and demonstrating sensitivity to capsaicin and various chemical agents. Upon activation, TRPV1 facilitates the influx of sodium and calcium ions into the cell, leading to neuronal depolarization and ultimately triggering signal transmission^[6]^.

TRPV1 is primarily expressed in neurons of the dorsal root ganglia and trigeminal ganglia^[6]^.Research indicates that TRPV1 is a pivotal molecule in regulating nociception, particularly thermal hyperalgesia following inflammation. Experimental evidence confirms that under inflammatory conditions, TRPV1 activation is a crucial mechanism for the induction of thermal hyperalgesia. This evidence is supported by the effectiveness of genetic or pharmacological suppression of TRPV1 in reducing or blocking inflammatory thermal hyperalgesia, thereby endorsing the potential of TRPV1 as a therapeutic target for treating inflammation-related pain^[8]^.

The pathogenesis and progression of allergic diseases involve a complex immune response, engaging a multitude of cells and molecules. These entities secrete specific cytokines during allergic reactions, which further escalate IgE production and the inflammatory response. Current research is increasingly focused on elucidating the neuro-immunological mechanisms underlying allergic rhinitis^[9]^.The fundamental premise for vidian neurectomy lies in the bidirectional regulatory interplay between the neural and immune systems^[10]^.In discussing TRPV1 expression at sensory nerve terminals and its role in airway inflammation, the concept of the naso-brain axis becomes increasingly apparent^[11]^. This emphasizes how the sensory nervous system can convey peripheral signals directly to the central nervous system, thereby influencing the brain’s immune responses and inflammation regulation. This concept not only broadens our understanding of neuro-immune interactions but also provides a novel perspective for exploring new therapeutic approaches to airway inflammation.An increasing body of research is focusing on the relationship between TRPV1 and allergic diseases, exploring how this ion channel may influence the immune dynamics underlying these conditions^[12]^.Studies indicate that TRPV1 mediates downstream inflammatory responses by modulating the activation of CD4+ T cells. TRPV1 activation interacts with the signaling pathways of these cells, influencing cytokine release and thus impacting the pathogenesis and progression of allergic rhinitis^[13]^.In research concerning TRPV1’s role in allergic asthma, TRPV1 may modulate the pathogenic mechanisms by regulating the functions of CD4+ T cells, thereby influencing the disease’s onset and progression^[14]^.In certain studies, TRPV1 is considered a therapeutic target for dermatological conditions such as allergic dermatitis. TRPV1 may contribute to the development of inflammatory skin diseases by modulating the release of inflammatory mediators, affecting epidermal barrier function, and regulating skin immune responses^[15]^.

Bibliometric analysis is an essential methodology for reviewing published articles, analyzing current research hotspots, and predicting future research trends^[16]^.Through the comprehensive utilization of bibliometric analysis, we can thoroughly assess research dynamics and trends within a specific field, identify key research institutions and scholars along with their contributions, thereby unveiling research hotspots and frontier areas^[17]^.Employing bibliometric techniques enables an intuitive examination of research articles related to TRPV1 and allergic diseases, uncovering research hotspots in this field over the past two decades. This approach provides a foundation for globally exploring the relationship between TRPV1 and allergic diseases, revealing the intricacies of neuroimmune inflammatory mechanisms. The data for this study, derived from the WoS database, is analyzed using the WoS online analysis platform, CiteSpace, the bibliometrix package in R, and Microsoft Excel. These tools are utilized to organize and describe the characteristics of the literature, along with performing citation, collaboration, and co-occurrence visual analyses, thereby illustrating the research hotspots and developmental trends in the study of TRPV1 and allergic diseases.

## MATERIALS AND METHODS

### Date Source

A thematic keyword field search was conducted in the Web of Science (WoS) Core Collection databases, specifically utilizing the SCI-EXPANDED and SSCI as sources.To enhance the precision and comprehensiveness of the search results, the query was standardized according to the MeSH (Medical Subject Headings) terminology, resulting in the final search string: “(ALL=(allergy OR allergies OR hypersensitivity OR “allergic reaction” OR “anaphylaxis” OR “Hypersensitivities” OR “Allergic Reactions”) AND ALL=(TRPV1 OR “transient receptor potential vanilloid 1” OR “capsaicin receptor” OR “vanilloid receptor 1”))”.This study completed its literature search on March 11, 2024, with the search timeline set from January 1, 2000, to December 31, 2023, ensuring a comprehensive collection of data. The document types were specified to include “Article, Review Article, Meeting Abstract, Proceeding Paper, Editorial Material, Early Access, Book Chapter, Letter, Correction, and News Item or Publication With Expression Of Concern” to broaden the inclusiveness of the results. Following the exclusion of irrelevant or duplicate publications, a total of 1,045 articles were identified through the search.

### Data Analysis

Utilizing the online analysis feature and citation report function of WoS, the search and data analysis were conducted on annual publication volume and other metrics. The data were organized into separate text documents based on categories such as year, authors, geographic distribution of countries and institutions, journals, and highly cited papers. During the information export, the option “Full Record and Cited References” was selected. The results were then imported into Excel for statistical analysis and graphing, enabling the exploration of trends and hotspots in TRPV1 and allergic disease research from 2000 to 2023.

The retrieved literature was exported in txt format and saved under the name “download_**.txt,” where ** represents the serial number of the exported documents. Subsequently, using CiteSpace 6.2.R4 and the bibliometrix package in R, the literature was imported for further visualization analysis. This process involved visualizing data on country and regional distribution, publishing institutions, journals, high-frequency keyword co-occurrence knowledge maps, keyword clustering, and burst detection. The objective was to investigate the relationship between TRPV1 and allergic diseases and to identify research hotspots and cutting-edge advancements in the field.

## RESULTS

### General Data and Annual Output

From 2000 to 2023, a total of 1,045 articles related to TRPV1 and allergic diseases were retrieved, categorized into 11 different types of publications. Among these, original research articles constituted approximately 81% (Table 1), indicating that research on TRPV1 and allergic diseases is predominantly focused on original studies. As illustrated in Figure 1, research on TRPV1 and allergic diseases was relatively scarce prior to 2008. Subsequently, the global publication volume on TRPV1 and allergic diseases demonstrated modest annual fluctuations with occasional declines, yet overall exhibited a stable growth trajectory. The highest number of publications was recorded in 2020, totaling 70 articles. Based on this data, 2020 emerges as a significant year during which research exploring the relationship between TRPV1 and allergic diseases peaked in popularity. This aligns with the increasing attention being accorded to TRP receptors in the scientific community.

**Table 1.**
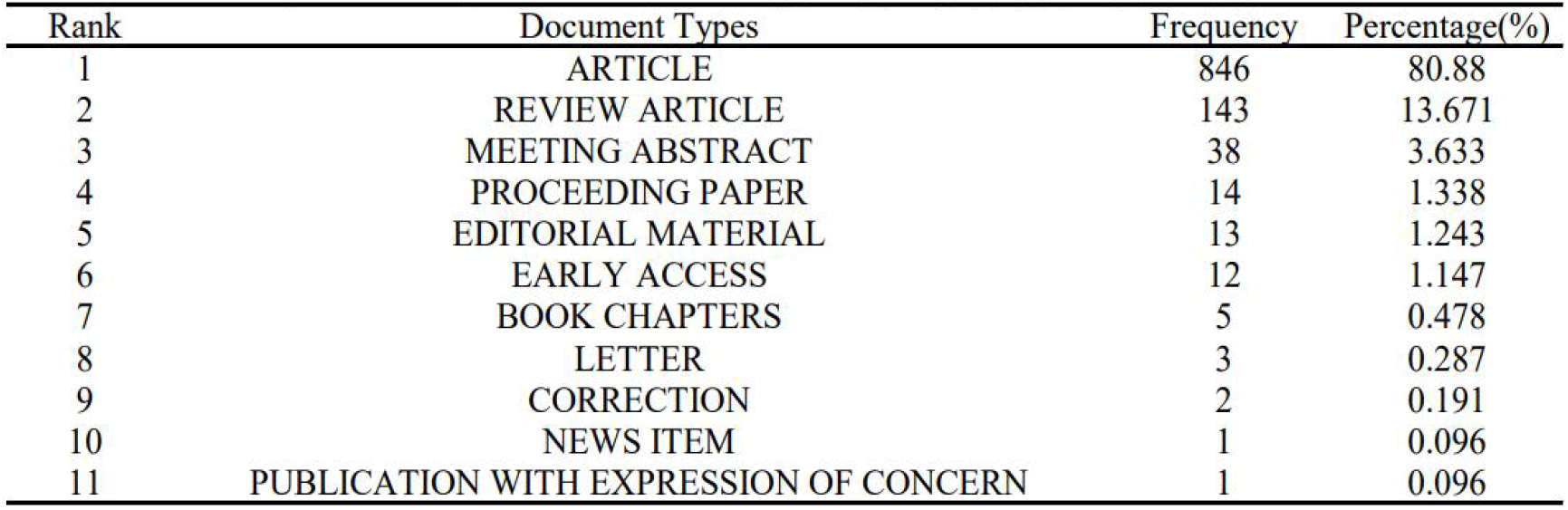
Quantities of Different Types of Articles.

**Figure 1.**
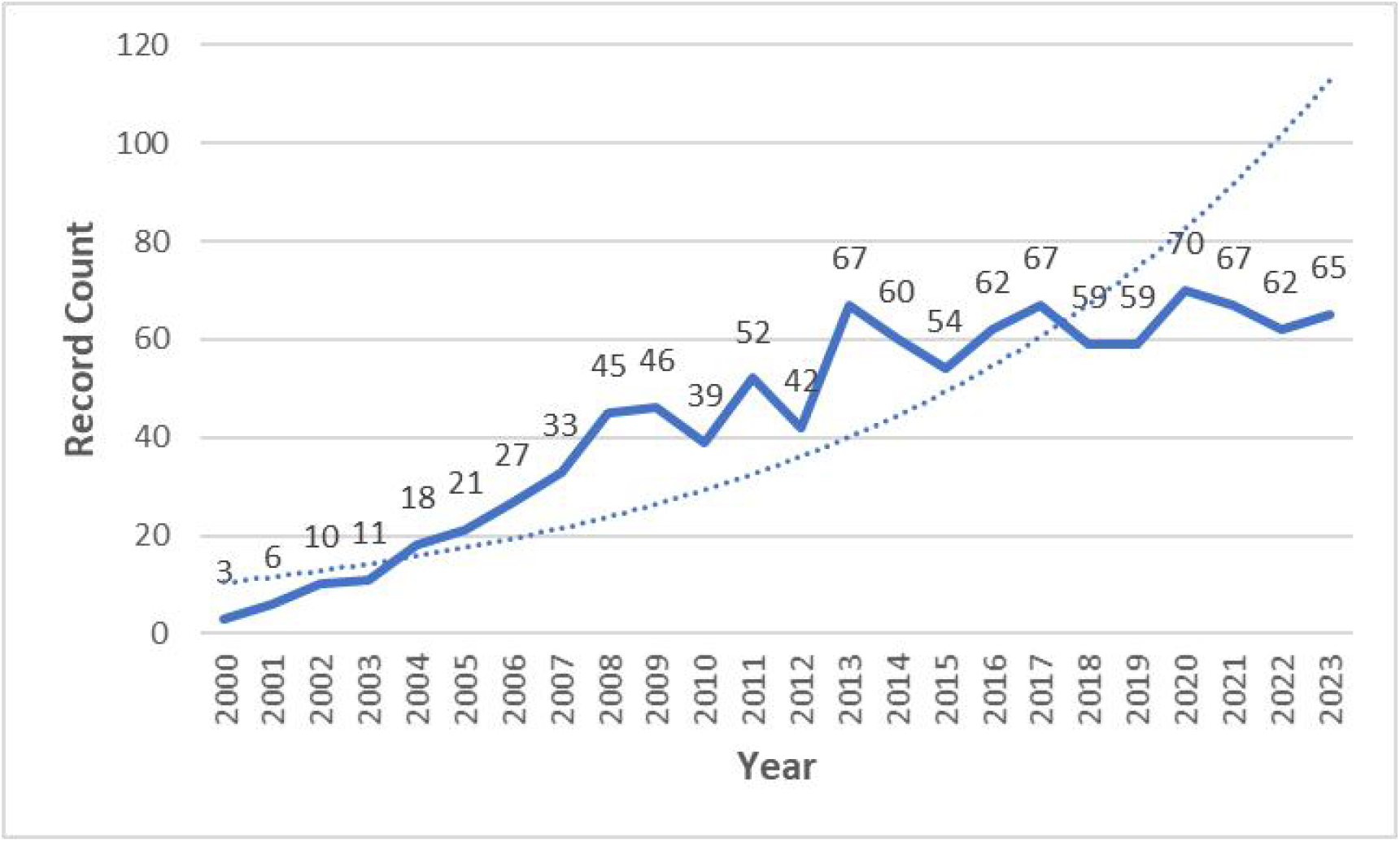
Annual Number of Published Research Articles.

### Distribution of Countries and Institutions

In this study, an analysis of 1,045 related articles in the database reveals that researchers from 57 countries and regions have conducted studies in this field. The publication volumes for the top 10 countries are presented in Table 2. The United States has the highest number of published articles (400 articles, accounting for 38.28%), followed by China (164 articles, accounting for 15.69%), and the United Kingdom (108 articles, accounting for 10.34%). Although China began its research later, it has quickly advanced to the forefront, now leading globally in the annual number of publications on this topic. This indicates that China is increasingly prioritizing research on the relationship between TRP receptor channels and allergic diseases.Figure 2A presents a world map illustrating the distribution density of countries that have published research articles on TRPV1 and allergic diseases from 2000 to 2023. The number of related papers published by research institutions to some extent reflects their innovation and competitiveness in this field. Between 2000 and 2023, a total of 1,179 institutions worldwide published articles on TRPV1 and allergic disease research. The institution with the highest number of publications is Johns Hopkins University (USA, with 56 articles, accounting for 5.36%). This institution primarily focuses on research related to the function of pain-related genes in nociceptive neurons. Among the top 10 institutions in terms of publication volume, eight are from the United States. This study has compiled a list of the top 21 institutions by publication volume, as detailed in Table 3. These research institutions have made outstanding contributions to the study of TRPV1 and allergic diseases.

**Table 2.** Top 10 global nations contributing to research outcomes.

| Rank | Country | Frequency | Percentage(%) |
| --- | --- | --- | --- |
| 1 | USA | 400 | 38.28 |
| 2 | PEOPLES R CHINA | 164 | 15.69 |
| 3 | ENGLAND | 108 | 10.34 |
| 4 | JAPAN | 101 | 9.67 |
| 5 | GERMANY | 99 | 9.47 |
| 6 | ITALY | 52 | 4.98 |
| 7 | SOUTH KOREA | 45 | 4.31 |
| 8 | CANADA | 43 | 4.12 |
| 9 | FRANCE | 35 | 3.35 |
| 10 | BELGIUM | 33 | 3.16 |

**Table 3.** Number of publications by global research institutions.

| Rank | Institutions | Country | Frequency | Percentage(%) |
| --- | --- | --- | --- | --- |
| 1 | JOHNS HOPKINS UNIVERSITY | USA | 56 | 5.36 |
| 2 | UNIVERSITY OF LONDON | UK | 51 | 4.88 |
| 3 | UNIVERSITY OF TEXAS SYSTEM | USA | 43 | 4.12 |
| 4 | HARVARD UNIVERSITY | USA | 42 | 4.02 |
| 5 | IMPERIAL COLLEGE LONDON | UK | 34 | 3.25 |
| 6 | JOHNS HOPKINS MEDICINE | USA | 34 | 3.25 |
| 7 | PENNSYLVANIA COMMONWEALTH SYSTEM OF HIGHER EDUCATION PCSHE | USA | 34 | 3.25 |
| 8 | HARVARD MEDICAL SCHOOL | USA | 33 | 3.16 |
| 9 | UNIVERSITY OF CALIFORNIA SYSTEM | USA | 32 | 3.06 |
| 10 | UNIVERSITY OF PITTSBURGH | USA | 30 | 2.87 |
| 11 | FREE UNIVERSITY OF BERLIN | Germany | 23 | 2.20 |
| 12 | GLAXOSMITHKLINE | UK (Headquarters) | 23 | 2.20 |
| 13 | QUEEN MARY UNIVERSITY LONDON | UK | 23 | 2.20 |
| 14 | KU LEUVEN | Belgium | 22 | 2.11 |
| 15 | SEOUL NATIONAL UNIVERSITY SNU | South Korea | 22 | 2.11 |
| 16 | CHARITE UNIVERSITATSMEDIZIN BERLIN | Germany | 20 | 1.91 |
| 17 | HUMBOLDT UNIVERSITY OF BERLIN | Germany | 20 | 1.91 |
| 18 | UNIVERSITY OF CALIFORNIA SAN FRANCISCO | USA | 20 | 1.91 |
| 19 | CENTRE NATIONAL DE LA RECHERCHE SCIENTIFIQUE CNRS | France | 19 | 1.82 |
| 20 | KING S COLLEGE LONDON | UK | 19 | 1.82 |
| 21 | UNIVERSITY SYSTEM OF OHIO | USA | 19 | 1.82 |

**Figure 2.**
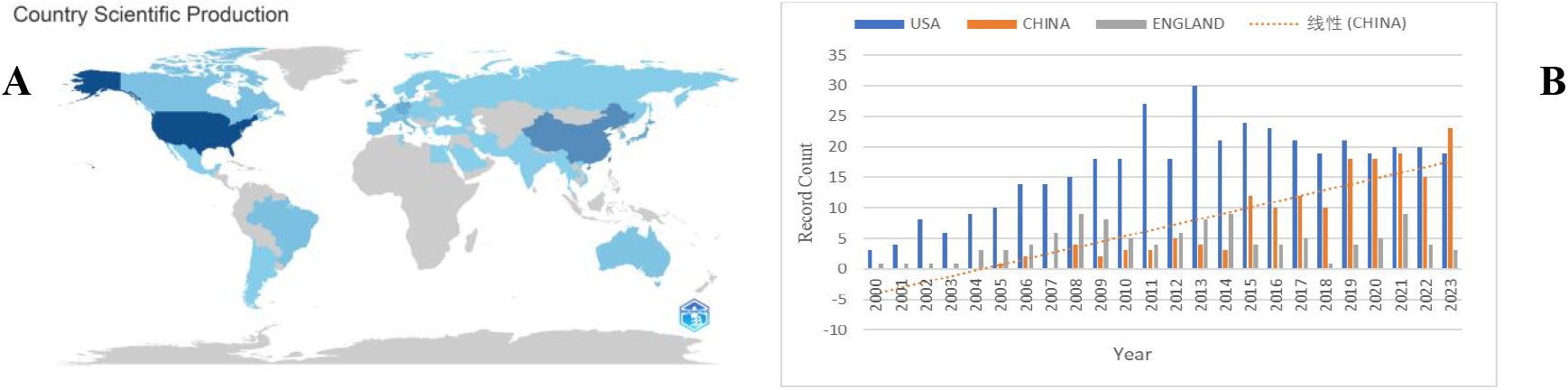
**A**:National distribution density map;**B**:The annual number of related papers published by the top three countries in terms of publication volume.

### Global Distribution of Authors and Analysis of Highly Cited Articles

Between 2000 and 2023, a total of 5795 authors published literature related to TRPV1 and allergic diseases. Table 4 lists the top five authors who have made the most significant contributions to the volume of publications in this study.The H-index is a quantitative metric that indicates a researcher has published H papers, each of which has been cited at least H times, while the citation count for their other papers does not exceed H^[18]^.The H-index is used to assess the quantity and level of an academic’s output, indirectly evaluating an individual’s research capabilities. The top publisher is Bradley Joel Undem from Johns Hopkins University in the USA, with 30 articles and an H-index of 59. His research spans physiology, respiratory systems, and neuroscience.The literature included in this study has been cited a total of 53,791 times, with the annual cumulative citation count of the published articles showing an increasing trend, as illustrated in Figure 3. This dataset includes three highly cited research papers related to the study, which are available online via the Web of Science (WoS) website, as shown in Table 5.Among them, the paper authored by Ferda Cevikbas et al., titled “A sensory neuron-expressed IL-31 receptor mediates T helper cell-dependent itch: Involvement of TRPV1 and TRPA1,” describes how TRPV1 targets and regulates itching, including conditions such as atopic dermatitis^[19]^.

**Table 4.** The top five authors who have made the most significant contributions to this study.

| Researcher | Frequency | Percentage(%) | Organizations | H-Index |
| --- | --- | --- | --- | --- |
| Undem, Bradley Joel | 30 | 2.871 | Johns Hopkins University | 59 |
| Anand, Praveen | 18 | 1.722 | Harvard University | 53 |
| Kollarik, Marian | 17 | 1.627 | University of South Florida | 32 |
| Woolf, Clifford | 16 | 1.531 | Harvard University | 136 |
| Ji, Ru-Rong | 13 | 1.244 | Duke University | 97 |

**Table 5.**
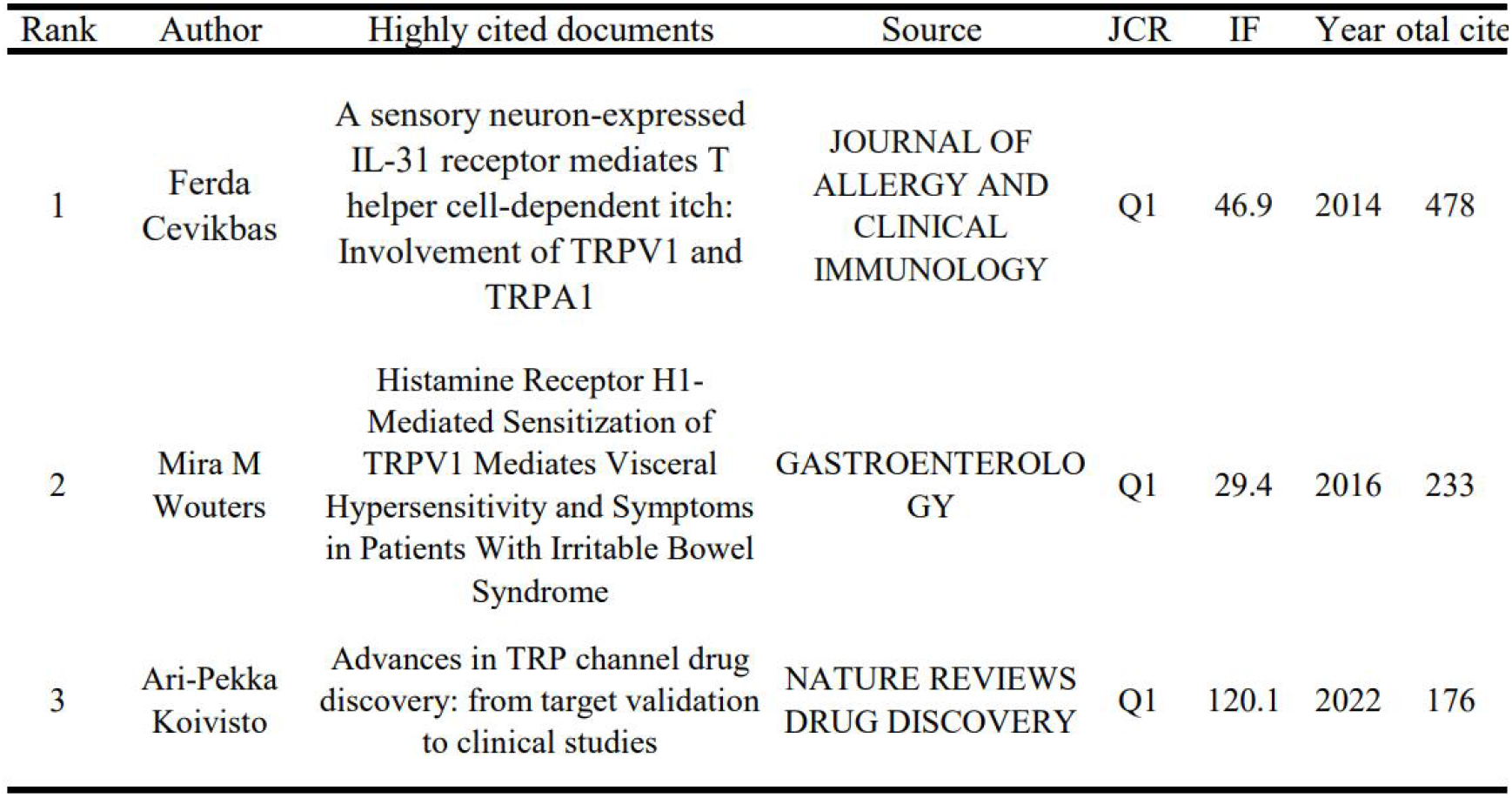
Highly cited papers.

**Figure 3.**
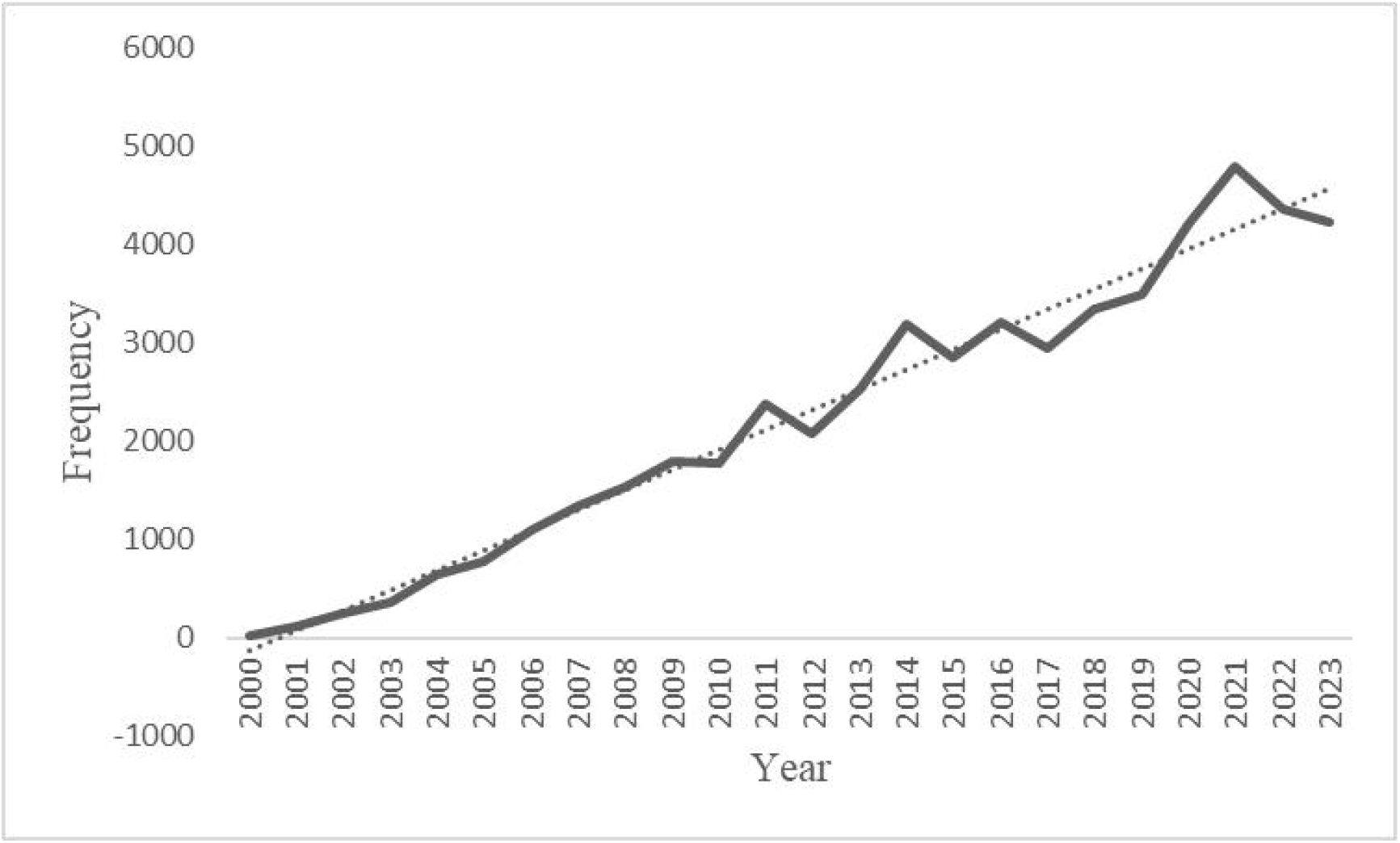
The annual citation frequency of research literature.

### Journals and Dual-overlay map of journals

Between 2000 and 2023, a total of 369 journals published research on TRPV1 and allergic diseases, with the top 10 journals listed in Table 6. These journals have each published 16 or more articles in this field. “PAIN,” with an Impact Factor (IF) of 7.4 in 2024, has published the most research articles (45 papers) and has an H-index of 26.It is noteworthy that among the top ten journals by publication volume, “GASTROENTEROLOGY” has the highest Impact Factor (IF = 29.4 in 2024) with 16 papers, yet its H-index is only 7. This indicates that the Impact Factor of a journal does not necessarily reflect its stature in the research field. It suggests that the IF, H-index, and citation frequency of the journal should be considered comprehensively.

**Table 6.** Journal Publication Analysis.

| Publication Titles | Frequency | Times Cited | Average per item | IF | JCR | H-Index |
| --- | --- | --- | --- | --- | --- | --- |
| PAIN | 45 | 1981 | 44 | 7.4 | Q1 | 26 |
| JOURNAL OF NEUROSCIENCE | 40 | 4487 | 112.18 | 5.3 | Q1 | 32 |
| NEUROGASTROENTEROLOGY AND MOTILITY | 35 | 1082 | 30.91 | 3.5 | Q2 | 19 |
| MOLECULAR PAIN | 33 | 1680 | 50.91 | 3.3 | Q3 | 21 |
| PLOS ONE | 23 | 873 | 37.96 | 3.7 | Q2 | 15 |
| JOURNAL OF ALLERGY AND CLINICAL IMMUNOLOGY | 22 | 1036 | 47.09 | 14.2 | Q1 | 11 |
| AMERICAN JOURNAL OF PHYSIOLOGY GASTROINTESTINAL AND LIVER PHYSIOLOGY | 16 | 603 | 37.69 | 4.5 | Q1 | 13 |
| GASTROENTEROLOGY | 16 | 750 | 46.88 | 29.4 | Q1 | 7 |
| NEUROSCIENCE | 16 | 713 | 44.56 | 3.3 | Q3 | 14 |
| SCIENTIFIC REPORTS | 16 | 257 | 16.06 | 4.6 | Q2 | 8 |

The dual-overlay map of the journal shows citing journals on the left and cited journals on the right. The curved lines between the left and right sections of the map represent citation links. The z-Scores function highlights stronger, more coherent trajectories, with higher scores represented by thicker connecting lines. Figure 4 illustrates that the fields of molecular biology, biology, immunology (yellow trajectory, z=7.88, f=90,588) and medicine, healthcare, clinical sciences (green trajectory, z=2.57, f=31,428) are likely to be influenced by publications in molecular biology, biology, and genetics. A substantial number of papers are distributed in the domains of molecular biology, biology, and immunology, with some in medicine, healthcare, and clinical sciences.

**Figure 4.**
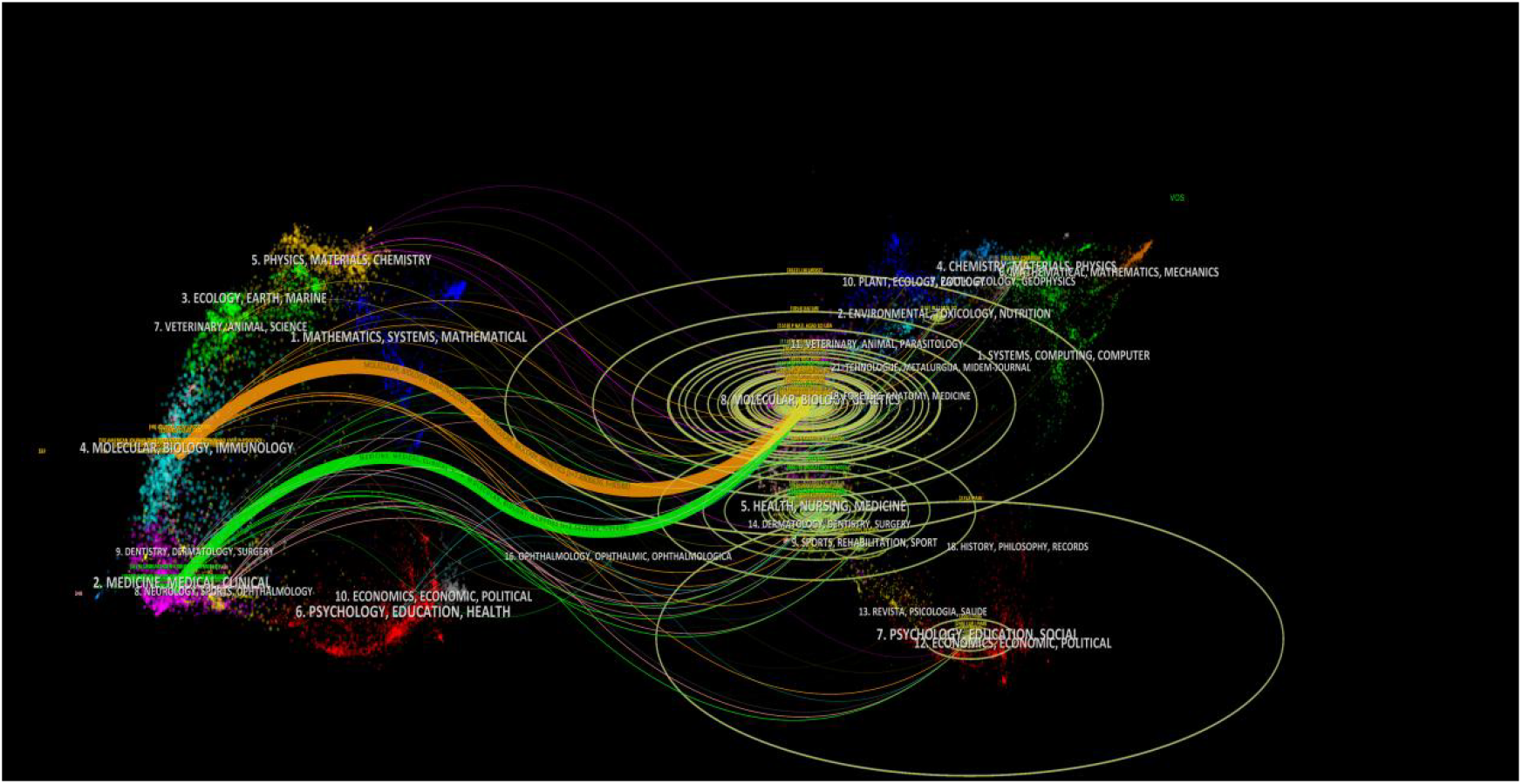
Dual-overlay map of the journal.

### Author Collaboration and Institutional Interaction Network Diagram

Using the CiteSpace 6.2.R4 tool with article authors as nodes for parameter configuration, a visualization illustrating the collaboration relationships among authors of all research articles was created, as seen in Figure 5A. As shown in the figure, a network comprising 615 nodes and 1,131 links is displayed, with a density of 0.006, illustrating visible collaboration among them. The analysis results reveal that Bradley Joel Undem is the author with the highest number of publications and leads the largest research team, having published numerous papers related to this field in recent years.The institutional collaboration relationships are shown in Figure 5B, where the institution with the highest centrality is Johns Hopkins University (centrality of 0.18), followed by the University of London (centrality of 0.13), with the University of Texas System ranking third (centrality of 0.12). Researchers have independently formed multiple collaborative networks, with each institution exhibiting close cooperative relationships.

**Figure 5.**
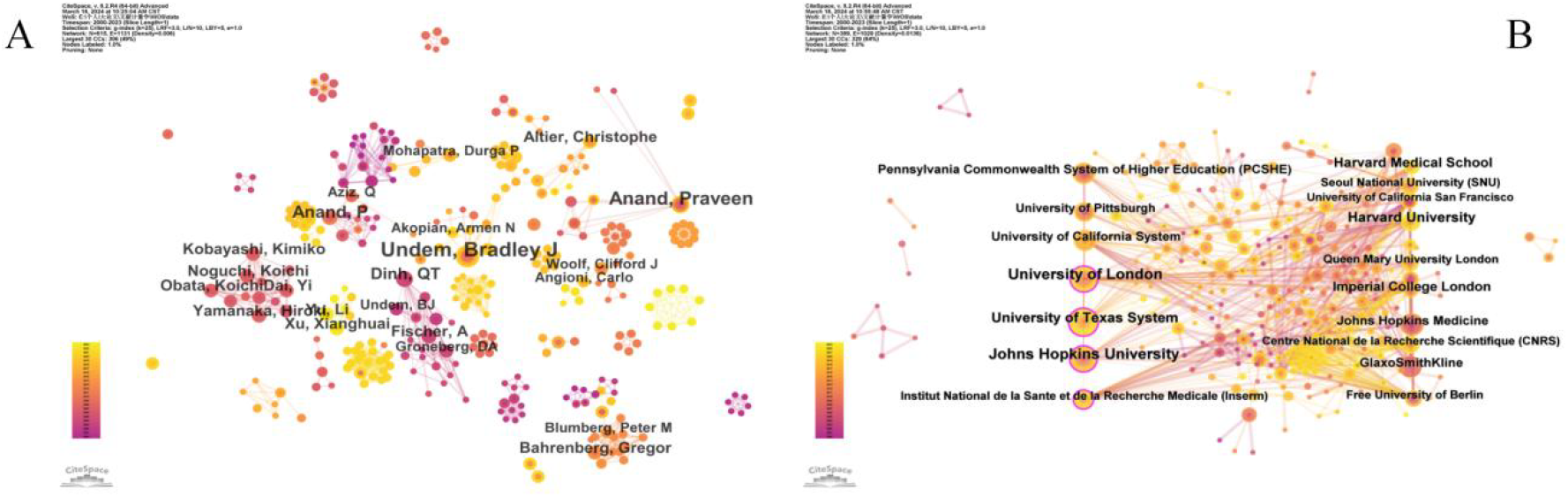
**A:**Author Collaboration Network Diagram; **B:**Institutional Interaction Network Diagram

### National or Regional Collaboration Network Diagram

According to the visualization results from previous studies using WoS, a total of 57 countries and regions worldwide have participated in research related to TRPV1 and allergic diseases. The collaboration map is depicted in Figure 6A. Using the CiteSpace 6.2.R4 software, we obtained a network with 57 nodes and 344 links, with a density of 0.2155. The top five countries in terms of centrality are the United States (centrality: 0.32) and Germany (centrality: 0.21), with Japan, South Korea, and France tied for third place with a centrality of 0.13. This indicates that the top five countries are all developed nations, highlighting the close collaboration among these countries.An analysis of the cooperation relationships between countries was conducted using a bibliometric online analysis platform, as shown in Figure 6B. The diagram illustrates that the United States and Germany engage in frequent collaborations with other countries, whereas China’s international collaborations are relatively less frequent compared to these nations. The research findings indicate that researchers from the United States play a leading role in this field and collaborate most frequently with scholars from other countries.The global contribution to research collaboration on TRPV1 and allergic diseases from 2000 to 2023 is depicted in Figure 6C. The Three-Field Plot is a visualization tool used in bibliometric analysis to display the main contents and interrelationships among three different domains, such as authors, journals, and countries. This type of diagram is typically presented in the form of a Sankey diagram, which effectively depicts the connections between elements from different domains, illustrating the flow and relationship among them^[20]^.The Sankey diagram obtained from this study, as shown in Figure 6D, indicates that the majority of the institutions are located in the United States, followed by China and Japan, with “PAIN” being the journal with the highest number of submissions.

**Figure 6.**
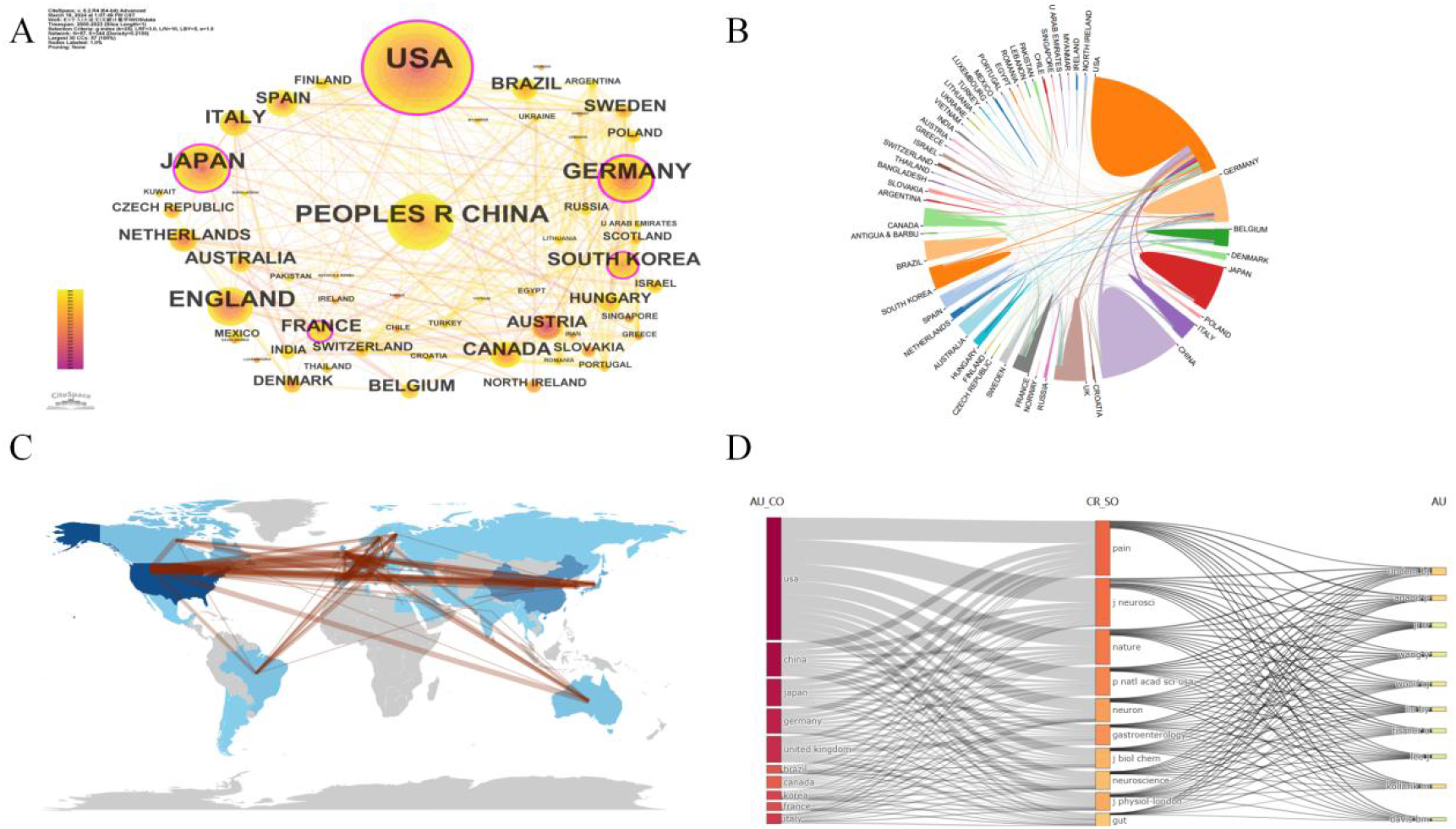
**A:**National or Regional Collaboration Network Map;**B:**Inter-country Collaboration Relationship Diagram;**C:**World Map of International Cooperation Between Countries;**D:**Sankey Diagram

### Keyword co-occurrence and burst

Keyword analysis indicates a primary focus on terms such as capsaicin receptor, TRPV1, sensory neurons, dorsal root ganglion, sensitivity, ion channel, expression, vanilloid receptor, substance-P, and TRP channels, as depicted in Figure 7A. The research directions are bifurcated into two main categories: studies on capsaicin receptors and activation research, illustrated in Figure 7B. The keyword themes are classified into eight major categories (Figure 7C). Timeline diagrams are frequently utilized to depict the number of published papers or trends in research within a specific field. The timeline diagram derived from keyword analysis in this study is presented in Figure 7D. Keyword bursts can unveil shifts in hotspots, emerging trends, or technological advancements in a research domain. As shown in Figures 8A and 8B, 49 keywords with high burst citation scores in the research area are highlighted, indicating recent research hotspots over the last five years, including mouse models, activation, contributes, mechanisms, model, allodynia, sensitization, TRPV1, oxidative stress, involvement, receptors, management, and atopic dermatitis.

**Figure 7.**
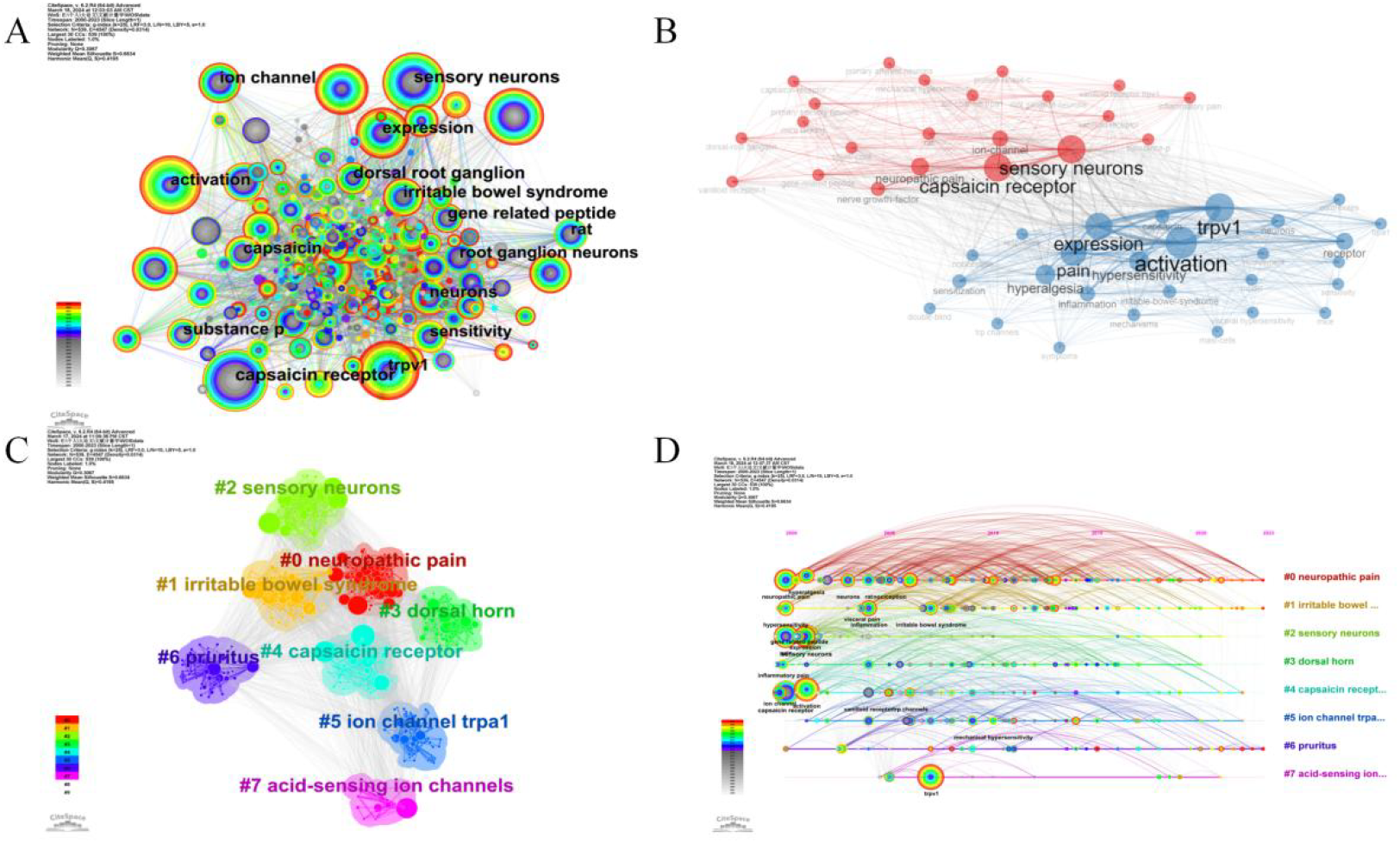
**A:**Keyword Co-occurrence Diagram;**B:**Keyword Research Direction Visualization Analysis;**C:**Keyword Cluster Analysis;**D:**Keyword Clustering Visualization Diagram.

**Figures 8.**
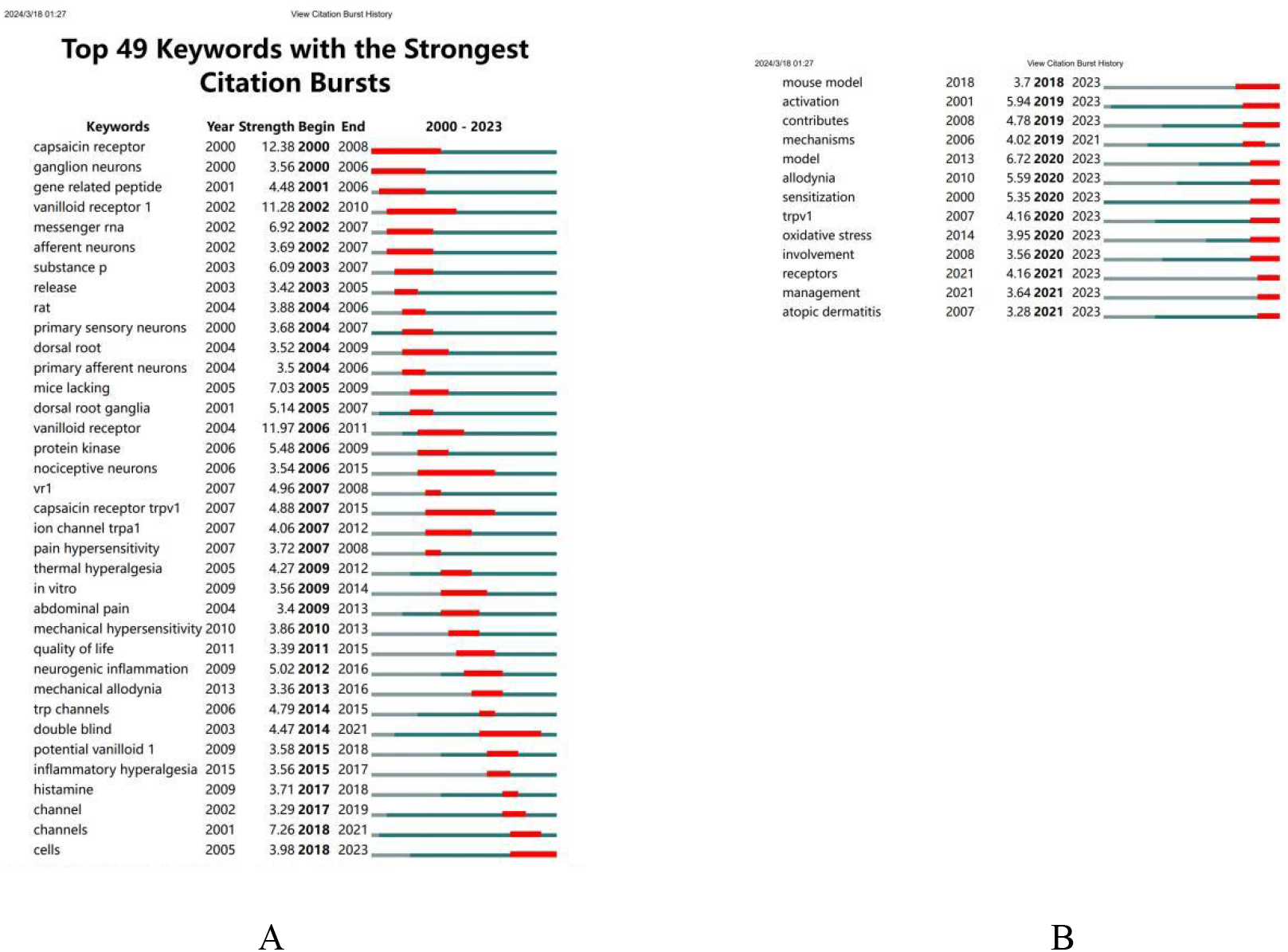
Keyword Burst Analysis Diagram.

## Discussion

Bibliometric analysis is a crucial tool for quantifying and evaluating academic publications, and it has been extensively applied across diverse research fields in recent years^[21]^. The publication trend of TRPV1 closely mirrors its research history: from the early 1970s to the late 1980s, research on TRPV1 was in its infancy, during which the receptor was gradually recognized and named. Subsequently, the receptor began to attract attention, leading to the emergence of related research publications, albeit in a limited and sporadic manner. Since the 1990s, there has been a continuous increase in the number of papers published in this field^[22]^. When not constrained by a specific time frame, results from the Web of Science indicate that research on the association between TRPV1 and allergic diseases began to emerge in the medical academic community starting from 1993^[23]^.In 1994, 1997, 1998, and 1999, there was one relevant paper published each year. However, in 2000, the number of publications related to this research increased exponentially. Consequently, this study sets the time frame from 2000 to 2023. According to the Web of Science database, the number of publications related to this research has shown a year-on-year increase, indicating a growing interest in this area within the academic community. This study elucidates the research focus and outcomes at different periods regarding TRPV1 and allergic diseases globally. It encompasses researchers, various countries and regions, publishing institutions, citing journals, and highly cited documents. By employing diverse visualization methods, analyses of collaborations among authors, interactions between countries, and interactions among publishing institutions are conducted to explore the global distribution related to this research area. Furthermore, through keyword clustering and burst analysis, the latest hotspots and trend directions associated with this research are examined.

### Analysis of the research strength of global researchers, countries and regions, publishing institutions, and publishing journals

From the perspective of national and regional distribution, the United States currently leads globally in this field in terms of both publication volume and international collaborations, holding an absolute advantage in scientific research cooperation. Following the U.S. in publication volume are China, the United Kingdom, Japan, and Germany. However, in terms of international research collaboration, the subsequent rankings are Germany, Japan, South Korea, and France. While China ranks second globally in publication volume, its international collaborations are relatively limited. Notably, China has experienced a surge in the annual number of publications. Despite its late start in research, China has gradually assumed a significant position in recent years. For instance, in 2023, China surpassed the United States in the number of publications, becoming the leading country in publication volume.

In this bibliometric analysis, a notable observation is the prominent standing of Johns Hopkins University on a global scale concerning publication volume in TRPV1 channels and allergic disease-related research. Among the top 21 institutions worldwide in publication volume, 10 are situated in the United States, 5 in the United Kingdom, 3 in Germany, and 1 each in Belgium, South Korea, and France, with no Chinese institutions represented. Particularly noteworthy is Johns Hopkins University’s leading position in publication volume, underscoring its pivotal role in advancing scientific knowledge in the realm of allergy-related research.The accomplishments of Johns Hopkins University in this domain likely derive from its comprehensive prowess in biomedical research, characterized by robust funding support, state-of-the-art laboratory infrastructure, and an environment conducive to attracting esteemed scientists and researchers. A thorough examination of the literature emanating from Johns Hopkins University reveals a breadth of research encompassing not only the fundamental biological roles of TRPV1 in allergic diseases but also extending to clinical applications. These endeavors encompass, among others, the exploration of novel treatments for conditions such as allergic asthma and allergic rhinitis^[24-25]^.The exemplary quality and innovation of these studies are evidenced by their high citation rates and widespread recognition within the academic community. Notably, Johns Hopkins University’s research achievements in this domain have exerted a significant influence on future research trajectories and methodological approaches. For instance, their investigations into the TRPV1 channel as a therapeutic target for allergic diseases not only provide fresh insights into the physiological mechanisms underlying these conditions but also lay the groundwork for the development of more precise and less adverse-effect-prone therapeutic interventions.

In the analysis of researchers, this study delineates the top five contributors based on publication volume, all hailing from the United States. An author interaction network analysis further highlights the foremost scholar in terms of relevance and publication output, identified as Bradley Joel Undem from Johns Hopkins University. A comprehensive examination of Professor Undem’s research endeavors unveils his substantial contributions, particularly pertaining to the intricate involvement of sensory nerves, notably vagal afferent fibers, in modulating airway inflammatory responses^[26]^. Through collaborations with experts in immunology, molecular biology, and other disciplines, he has emphasized the significance of interdisciplinary cooperation, resulting in publications widely cited within the academic community. Articles related to this research predominantly appear in journals specializing in pain studies and neuroscience, with the journal PAIN assuming a prominent position in publication volume concerning TRPV1 channels and allergic diseases. As a leading journal in the pain research domain, PAIN has consistently published high-quality research papers spanning molecular mechanisms to clinical applications.The most cited article in this research field is “A sensory neuron-expressed IL-31 receptor mediates T helper cell-dependent itch: Involvement of TRPV1 and TRPA1” by Ferda Cevikbas et al. This study delves into the role of TRPV1 and TRPA1 in mediating itch sensations dependent on T helper cells, particularly under the influence of the IL-31 receptor expressed in sensory neurons. By elucidating the involvement of IL-31 in sensory nerve regulation and the functions of TRPV1 and TRPA1 in itch transmission, it sheds light on potential novel targets for treating allergic rhinitis and associated itch, especially among patients unresponsive to traditional treatments. Modulation of the activity of these channels holds promise for offering innovative therapeutic strategies for allergic rhinitis^[19]^.

### Keyword analysis

The keyword visualization analysis study reveals that research on TRPV1 in allergic diseases primarily centers on key themes such as the capsaicin receptor, TRPV1, sensory neurons, dorsal root ganglion, sensitivity, ion channel, expression, vanilloid receptor, substance-P, and TRP channels. These themes delineate two main research directions: investigations into the capsaicin receptor and studies focusing on activation mechanisms. This observation underscores the pivotal role and central focus of TRPV1 in allergic disease research. Li F^[27]^ and colleagues delved into the role of TRPV1 in pain and itch, highlighting how TRPV1-expressing neurons interact with immune cells by releasing neuropeptides under pathological conditions. This interaction facilitates the transmission of pain and itch signals. Such insights offer a novel perspective on the involvement of TRPV1 in allergic diseases, notably conditions like allergic rhinitis, asthma, and atopic dermatitis. In these conditions, TRPV1 activation may be intricately linked to the release of inflammatory mediators, pain propagation, and itch sensation generation.

Keyword clustering analysis unveils eight major thematic directions in current research. Though distinct, these themes are interconnected, highlighting the pivotal role of TRPV1 in diverse physiological and pathological processes and its intricate regulatory network. As previously mentioned, investigations into TRPV1 underscore its substantial involvement in neuropathic pain and itch, elucidating the neurobiological underpinnings of pain and itch sensations and their potential as therapeutic targets^[27-28]^. Research on the TRPV1 in irritable bowel syndrome (IBS) has elucidated the intricate interaction between gastrointestinal physiology and the nervous system. This underscores the promising prospect of alleviating IBS symptoms by modulating TRPV1 activity.^[29-30]^.Research on sensory neurons and the dorsal horn underscores the pivotal role of TRPV1 in both the central and peripheral nervous systems, elucidating its significance in the transmission of pain and itch by sensory neurons, as well as its critical involvement in central processing within the dorsal horn. Furthermore, the investigative focus on acid-sensing ion channels delves into their function in sensing acidic environments and modulating responses under associated pathological states, thereby broadening our comprehension of TRPV1 and its analogous channels in sensory physiology and pain perception^[31]^.

The burst of keyword results underscores a growing reliance on studies employing mouse models, underscoring their indispensability as instrumental platforms for elucidating the role of TRPV1 in pain perception, inflammatory responses, and allergic reactions. Through the utilization of mouse models, researchers can replicate human allergic conditions within controlled settings, facilitating comprehensive investigations into the precise functionalities of TRPV1 and identifying potential therapeutic targets^[32]^. The frequent occurrence of the keyword “activation” in research highlights a significant focus on elucidating how the activation of TRPV1 can simulate pain and inflammatory responses within the pathological context of allergic diseases. This entails in-depth investigations into the mechanisms underlying TRPV1 channel activation and the subsequent effects of such activation on pain transmission pathways and inflammatory responses. A comprehensive understanding of these processes is paramount for the development of novel therapeutics aimed at alleviating allergic diseases^[27]^. The emergence of “allodynia” within the array of keyword mutations in TRPV1 research signifies a concentrated inquiry into the mechanisms underpinning pathological pain, particularly those states surpassing typical physiological responses. This encompasses persistent pain resultant from nerve damage or inflammation, wherein the activation and modulation of TRPV1 assume pivotal roles in conveying aberrant pain signals. This underscores the imperative for a comprehensive comprehension of TRPV1’s involvement in such processes to facilitate the development of therapeutic strategies tailored to address neuropathic pain states^[33]^. The notable prevalence of “sensitization” in the analysis of keyword mutations within TRPV1 research underscores a keen research focus on the heightened sensitivity of TRPV1 in pain and inflammation pathways. This encompasses TRPV1’s enhanced responsiveness to pain signals following repeated or sustained stimulation, thereby accentuating the potential significance of TRPV1 channels in the onset of chronic pain and allergic conditions. The modulation of TRPV1 sensitization presents a promising avenue for the development of novel therapeutic interventions targeting associated ailments^[34]^. The inclusion of “oxidative stress” in the array of keyword mutations within TRPV1 research highlights the involvement of TRPV1 under conditions of oxidative stress and its implications for pain perception and the advancement of allergic diseases. Oxidative stress denotes an imbalance between free radicals and antioxidants in the body, which can trigger the activation of TRPV1 channels, subsequently culminating in heightened pain sensitivity and inflammatory reactions^[35]^. Indeed, the revelation underscores the therapeutic potential of controlling oxidative stress levels and modulating TRPV1 activity in mitigating pain and inflammatory responses precipitated by oxidative stress. By targeting these pathways, novel therapeutic approaches may hold promise in ameliorating symptoms associated with oxidative stress-related pain and inflammation^[36]^. The emergence of “atopic dermatitis” in the cluster of keyword mutations within TRPV1 research underscores a dedicated examination of the pathological mechanisms underlying skin diseases. Through discerning the role of TRPV1 in atopic dermatitis, recognition extends to the potential involvement of TRPV1 in analogous allergic conditions, like allergic rhinitis. This highlights the imperative of investigating the TRPV1 pathway to advance novel treatments for atopic dermatitis, while also offering insights into the management of other allergic diseases such as allergic rhinitis.

### Strength and Limitations

This study integrates the use of Citespace software, the bibliometrix package in the R language, and online literature analysis platforms to delineate the focal areas and directional trends of TRPV1 research within allergic diseases from 2000 to 2023. By employing bibliometric methodologies to quantify scientific literature, this approach offers the advantage of systematically organizing and unveiling trends, hotspots, and knowledge structures within the research domain. Such analyses aid in identifying research lacunae and cutting-edge topics, rendering them suitable for research assessment within big data environments. Furthermore, this methodology proves effective in tracking scientific advancements and technological innovations. The findings presented in this paper are poised to provide pertinent researchers and institutions with deeper insights into prevailing developmental trajectories, thereby fostering the advancement and application of research outcomes within the realm of TRP ion channels and allergic research.

The primary limitation of this paper pertains to its relatively confined data source, as it neglects to amalgamate information from alternative databases for a more comprehensive analysis. The study’s failure to aggregate collected data with information sourced from different databases such as PubMed precludes a holistic examination. Secondly, its emphasis on quantitative analysis may potentially overshadow the importance of assessing the quality and depth of the literature. Thirdly, the study relies solely on data extracted from the WoS database. Although WoS is consistently esteemed as a premier source for bibliometric analysis and visualization, limitations intrinsic to both the database and search strategy may result in incomplete coverage of pertinent literature. Nonetheless, the data obtained are objective and expansive, offering a representation of the current status and trends within the study’s scope. Additionally, literature published proximate to the search period may exhibit lower citation frequencies owing to its recent dissemination, potentially introducing biases in the literature’s quality assessment. Indeed, while lacking the depth characteristic of basic research, the quantitative methodology utilized offers an intuitive portrayal of prevailing viewpoints and emerging trends within the field. Despite these acknowledged limitations, it is undeniable that bibliometric methodologies can serve as a guiding compass for delineating directions in both basic and clinical research, thereby elevating research standards within the domain of TRP and allergic diseases.

## Conclusion

Through bibliometric and visualization analysis of the Web of Science dataset encompassing TRPV1 and allergic diseases from 2000 to 2023, a discernible trend emerges: research interest in this domain has steadily burgeoned over the years, with American researchers assuming a predominant role in its advancement. Numerous pertinent studies have surfaced in high-caliber journals. While earlier attention primarily centered on the receptor itself, there is now a pronounced shift towards establishing animal models and unraveling activation mechanisms. Future investigations are anticipated to delve deeper into areas such as sensitization mechanisms and receptor expression. Given its potential as a therapeutic target for allergic diseases, continued exploration of TRPV1 is imperative.

## Data Availability

All relevant data are within the manuscript and its Supporting Information files.

